# Integrated DNA estimation in tissue biopsy and detection in liquid biopsy by HBV-targeted NGS assay

**DOI:** 10.1101/2024.12.04.24318256

**Authors:** Hsin-Ni Liu, Selena Y. Lin, Ricardo Ramirez, Shin-En Chen, Zach Heimer, Roman Kubas, Fwu-Shan Shieh, Elena S. Kim, Yuanjie Liu, Daryl T.Y. Lau, Ting-Tsung Chang, Haitao Guo, Zhili Wang, Ying-Hsiu Su

**Affiliations:** The Baruch S. Blumberg Research Institute, Doylestown, PA, 18902, USA; JBS Science, Inc., Doylestown, PA,18902, USA; Gilead Sciences, Inc, Foster City, CA, 94404, USA; Department of Microbiology and Immunology; Cancer Virology Program, UPMC Hillman Cancer Center, University of Pittsburgh School of Medicine, Pittsburgh, PA, 15213, USA; Liver Center, Department of Medicine, Beth Israel Deaconess Medical Center, Harvard Medical School, Boston, MA, 02215, USA; Department of Internal Medicine, National Cheng Kung University Medical College, Center of infectious Disease and Signaling Research, National Cheng Kung University, Tainan, Taiwan, Republic of China

**Keywords:** HBV integration, HBV NGS, liquid biopsy, antiviral therapy monitoring, HBV-host junction sequences, HBV genetics

## Abstract

**Background & Aims:** Integrated HBV DNA (iDNA) plays a critical role in HBV pathogenesis, particularly in predicting treatment response and HCC. This study aimed to use an HBV hybridization-capture next-generation sequencing (HBV-NGS) assay to detect HBV-host junction sequences (HBV-JS) in a sensitive nonbiased manner to detect and estimate the iDNA fraction in tissue biopsies and HBV genetics by liquid biopsy.

**Methods:** HBV DNA from plasmid monomers, HBV-HCC cell line (SNU398, Hep3B, and PLC/PRF/5), tissue biopsies of patients with serum HBV DNA <4 log IU/ml, and matched urine and plasma of HBV patients were assessed by HBV-NGS. Junction-specific qPCR (JS-qPCR) assays were developed to quantify abundant HBV-JS.

**Results:** We demonstrated high coverage uniformity, reproducibility across all HBV genotypes A-D, and 0.1% sensitivity for detecting iDNA by the HBV-NGS assay. The sequence and structures of iDNA molecules from SNU398 and Hep3B are reported. An iDNA estimation model was developed using six abundant HBV-JS sequences identified from tissue biopsies by HBV-NGS assay and validated using total DNA of SNU398 and Hep3B cells. Furthermore, the utility of the HBV-NGS assay for HBV genetic analysis in liquid biopsies was explored using matched plasma-urine samples from three patients with serum HBV DNA levels ranging from high to undetectable. HBV-JS was detected in all body fluids tested, regardless of viral load.

**Conclusion:** These findings suggest that the iDNA fraction in tissue biopsies from patients with limited or undetectable serum HBV DNA can be estimated using a robust HBV-NGS assay, and a sensitive HBV genetics liquid biopsy can be obtained. This study highlights the potential of NGS-based methods to advance HBV management.

**Impact and implications:** This study applied an unbiased, sensitive, and robust HBV-captured NGS method to detect integrated HBV DNA (iDNA) in both tissue and liquid (blood and urine) biopsies and generated a model to estimate the intrahepatic iDNA quantity. The availability of this technology makes possible to not only estimate iDNA fraction in total HBV DNA in tissue biopsies, but also perform HBV genetics in a minimally invasive manner in chronic HBV patients for disease monitoring, HCC risk assessment, and for clinical research.

## Introduction

Integrated HBV DNA (iDNA) is found in almost all chronically infected HBV (CHB) patients [1, 2] and >85% HBV-associated HCC (HBV-HCC) [3]. iDNA is known to play a significant role in sustained HBsAg expression complicating antiviral therapy monitoring and in carcinogenesis [1, 4]. Although HBV DNA is much lower in HBeAg negative as compared to HBeAg positive CHB patients, the proportion of iDNA to total HBV DNA is much higher in HBeAg negative patients. Recently, the relative reduction of iDNA in liver tissue has been associated with antiviral therapy outcomes [4, 5] and reduced HCC risk [6]. Thus, the quantification and characterization of iDNA can be biomarker for monitoring antiviral therapy response, disease progression as well as assessing HCC risk.

Conventional PCR methods for iDNA detection, inverse nested PCR (invPCR) [7–11] or Alu-PCR [5, 12–15] are, inherently biased as they rely on Alu-sequences or restriction enzyme sites. Alu-PCR may focus on selecting HBV regions such as S, X, and C, while invPCR is biased toward the region of ∼1,400-1,800, including the DR1-2 region integration breakpoint hotspots.

This study applied the technological advanced hybridization-capture next generation sequencing (NGS) platforms to perform unbiased capture of the entire HBV genome with the primary objective to comprehensively detect and estimate integration breakpoints across the viral genome. We employed HBV-targeted NGS assay with the previously developed *ChimericSeq* [16] pipeline for detection of HBV-host junction sequence (HBV-JS) breakpoints in both tissue and liquid biopsy from CHB patients of a spectrum of serum HBV DNA levels. By using cell-line DNA and liver biopsy tissues from patients with serum DNA < 4 log IU/mL, we established a sensitive, specific, robust and unbiased method in detecting iDNA breakpoints and a linear model to estimate iDNA fraction. We reported the sequences and rearrangement of the one iDNA in SNU398 cell line and two major iDNA molecules in Hep3B cell line. The application of the HBV genetic liquid biopsy method and iDNA estimation model is executed and discussed.

## Materials and Methods

### Study Subjects

Four patient tissue biopsies are provided by the Hepatitis B Research Network, a research network. Three pairs plasma/urine liquid biopsy specimens were collected from Beth Israel Deaconess Medical Center (Boston, MA). Written informed consent was obtained and in accordance with the guidelines of Committee on Clinical Investigations (CCI) the Institutional Review Board for the Beth Israel Deaconess Medical Center. The CCI approved this study which was conducted in accordance with both the Declarations of Helsinki and Istanbul. All patients have a history of HBV infection. The clinical information is summarized in **Table 1** with listed Patient IDs that are blinded and not know to anyone outside the research group.

**Table 1.**
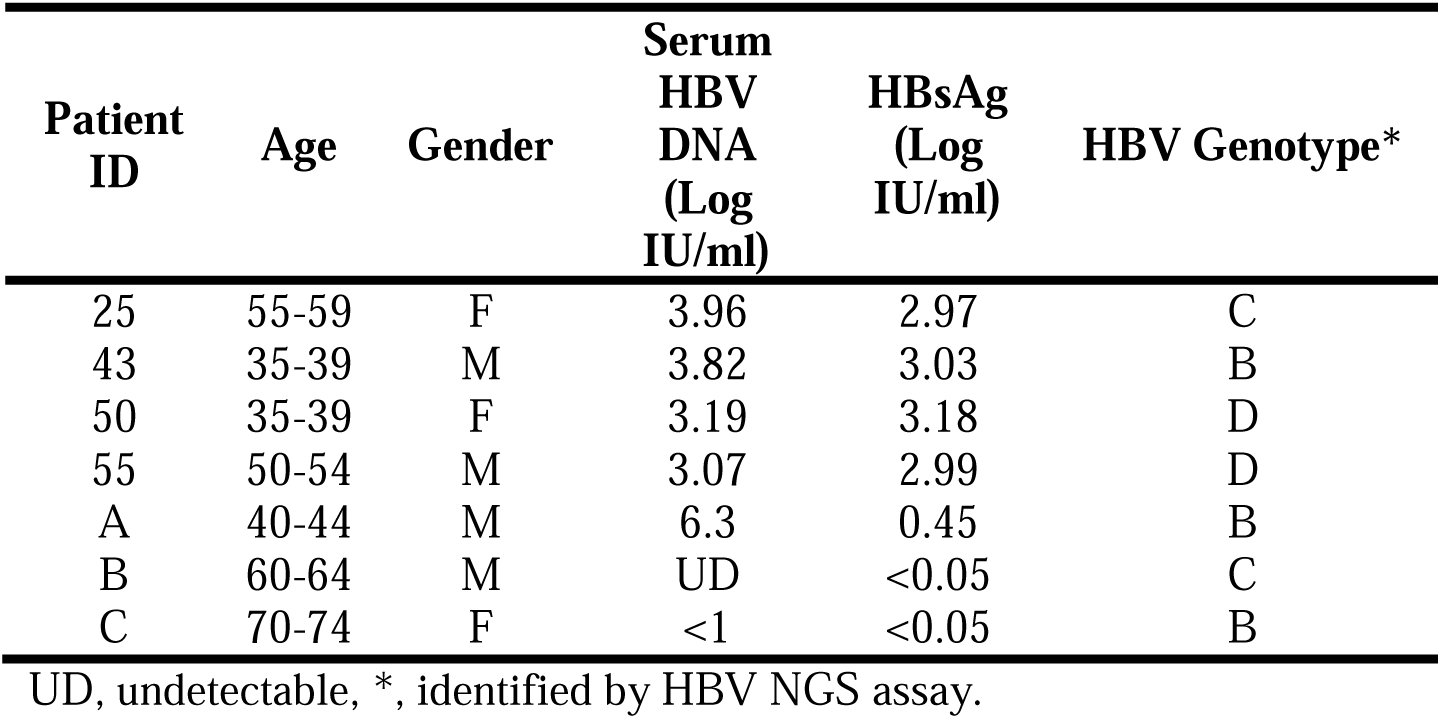
Study subjects.

### HBV plasmids and cell-line DNA controls

Plasmids containing a full-length monomeric HBV genome for genotypes A-D were synthesized by GENEWIZ, Inc (South Plainfield, NJ) and used for assay assessment. The plasmids contained a puC-GW-Kan vector backbone with HBV sequences from genotype A, B, C, and D with Genbank accession # EU054331.1, AB10428.1, GQ3581.58.1, and AF121240.1, respectively. DNA from three HBV-HCC cell-lines, SNU398, Hep3B, and PLC/PRF/5 [17–19] were used for HBV NGS assay characterization and iDNA estimation model validation.

### HBV-targeted NGS assay

Tissue biopsy DNA from four patients was extracted using AllPrep® DNA/RNA Micro Kit (Cat# 80284, Qiagen, Germantown, MD). For the initial tissue maceration, the samples were mounted into the β-mercaptoethanol-supplemented RLT lysis buffer and broken down in the PowerBead Tubes, filled with Ceramic 1.5 mm beads (Cat #13113-50, Qiagen) using Bead Mill 24 Homogenizer (Cat #15-340-163, Fisherbrand™, Pittsburgh, PA). Quantitative and qualitative analyses of isolated DNA were performed using NanoDropOne (Thermo Fisher Scientific, Waltham, MA). Approximately 100 ng of tissue DNA was fragmented (150 – 200 bp) by sonication and subjected to library DNA preparation using the xGen™ cfDNA & FFPE DNA library prep kit (Cat# 10010207, Integrated DNA Technologies, Coralville, Iowa), which contains unique molecular identifies (UMIs), according to manufacturer specifications. Cell-free DNA (cfDNA) from matched urine and plasma was extracted from three patients using the urine or plasma cell-free DNA isolation kit (Cat # 08872 and 08874, JBS Science Inc., Doylestown, PA). Plasma cfDNA amounts ranging from 3.7 – 7 ng and urine cfDNA amounts ranging from 35 – 75 ng was subjected to an HBV-targeted next generation sequencing (NGS) assay following the manufacturer’s instructions (JBS Science Inc.). Plasma and urine DNA libraries were subjected to one or two rounds of HBV hyb-capture, respectively. The captured library DNA was pooled for NGS analysis on the Illumina NovaSeq platform for 2 × 150 bp sequencing.

### Identification of HBV-JS and development of junction specific PCR assays

The sequencing data was analyzed using an Advanced *ChimericSeq bioinformatics pipeline* (JBS Science Inc.) upgraded from our previously developed software *ChimericSeq* [16]. The 42 HBV genomes described in McNaughton *et al.*, 2020 [20] were used as HBV references in sequence alignment. The *ChimericSeq pipeline* generates UMI consensus sequences and determines characteristics of HBV DNA in each sample, including genotype, % HBV reads of total NGS reads, % HBV-JS of all HBV reads, depths of coverage along the HBV genomes, HBV and host breakpoint coordinates, and the depths of individual HBV-JS. Selected HBV-JSs underwent junction-specific quantitative PCR (qPCR) assay development for quantitation of each HBV-JS by qPCR for iDNA estimation model development and validation. All the primers and assay information are detailed in **Supplementary Table 1.** For model validation, two HBV-JSs from cell-line SNU398, S1-GPR42 (hg38 Chr19:35376685, HBV nt. 448) and S2-LINC01531 (hg38 Chr19:35410237, HBV nt. 1301), and two most abundant HBV-JSs from Hep3B, H1-LINC00379 (hg38 Chr13: 91169310, HBV nt. 2056) and H2-ROBO2 (hg38 Chr3: 75917792, HBV nt. 1793), were used as detailed also in **Supplementary Table 1**. All the sequence alignment in the pipeline was done using bwa mem [21].

### Development of iDNA estimation model

For iDNA quantitation estimation using the *ChimericSeq* output, we used HBV-JS as a marker to estimate its fraction in total HBV reads (% HBV-JS read) and compared it to the % HBV-JS by HBV-JS specific qPCR assays. A linear model was developed using % HBV-JS reads as the independent variable and % HBV-JS by qPCR assays as dependent variable, as illustrated in **Supplementary Figure 1**. To generate the linear model for iDNA estimation using NGS data, we identified 6 HBV-JS with two criteria, at least 10% of total HBV-JS reads in the sample or can expand the applicable range (x-axis) of this model, from patients having less than 4 log IU/mL HBV serum viral load. Junction-specific qPCR for each of the selected junction sequences were developed, as detailed in **Supplementary Table 1**. The percentage of each HBV-JS in total HBV DNA (HBV CG1) quantified by HBV pol/S (246-309) PCR Quantification Kit (Cat# qPCR-013, JBS Science Inc.) was obtained. The percentage of each HBV-JS among total HBV genome copies (defined as “qPCR ratio”) in the sample was calculated and used as dependent variable (y-axis) using the calculation: 2(-ΔCp), where ΔCp = Cp(HBV-JS) - Cp(CG1). Likewise, the % of each HBV-JS by NGS reads among total HBV genome copies in a sample was calculated for each HBV-JS using NGS reads (defined as “NGS ratio”) and used as independent variable (x-axis) with the calculation: HBV-JS count*22/HBV read count of that sample, where 22 is the approximate number of 143-bp NGS reads that constitute a full length HBV (3.2 kb) assuming the size of iDNA is approximately a full-length. A linear model was built using function “lm” with formula y ∼ 0 + x and intercept set to 0 in R. Note the iDNA estimated by the linear model can be >100% if each integrated DNA is shorter than 3.2 kb because of deletion due to integration or resulted from rearrangement.

### Statistical analysis

To determine the relationships between HBV-JS quantified by qPCR and by NGS, Pearson’s correlation test was first used to obtain the correlation coefficient and the significance level (function “cor.test”). After finding a significant correlation between the two matrices, a linear regression model was built using the function “lm” and evaluated with the leave-one-out cross validation (LOOCV) method (R package “caret”). The adjusted-R^2^ and a p-value were reported for the regression model, and the root mean squared error (RMSE) and R^2^ values were reported for the assessment of the model performance. All statistical analyses were performed using R software version 4.1.2 (R Core Team, 2021). A p-value of 0.001 was considered statistically significant.

## Result

### Uniformity of HBV DNA capture and reproducibility of HBV-NGS assay assessed by full length HBV genomes

To determine the uniformity of HBV DNA capture and reproducibility of the commercially available HBV-NGS assay, plasmids containing a monomeric DNA genome of four HBV genotypes A – D were subjected to HBV-NGS assay in four replicates, detailed in Materials and Methods. HBV DNA coverage maps for each genotype are shown in **Figure 1**. Uniformity was evaluated by plotting the depth of HBV DNA coverage. As shown in **Figure 1A**, the HBV-probe panel was able to capture 100% of HBV genomic positions for all four genotypes tested. The coverage depths of every position except for both ends (50 nt on each end) are within a 2-fold difference relative to the mean coverage across the HBV genome (log2[coverage fold change] = [-1,1]). Regions near nt positions 850 and 2550 in genotype D had slightly less capture efficiency, but within a 2-fold difference of the mean coverage, compared to the rest of the genome. A slight decrease in capture efficiency at the end of the linear monomer adjacent to the plasmid vector backbone is expected, because of the reduced homology of the templates to the probe due to the shorter HBV DNA at the junction of the plasmid backbone. Nevertheless, the capture efficiency for all four genotypes was within a 2-fold difference relative to the mean coverage.

**Figure 1.**
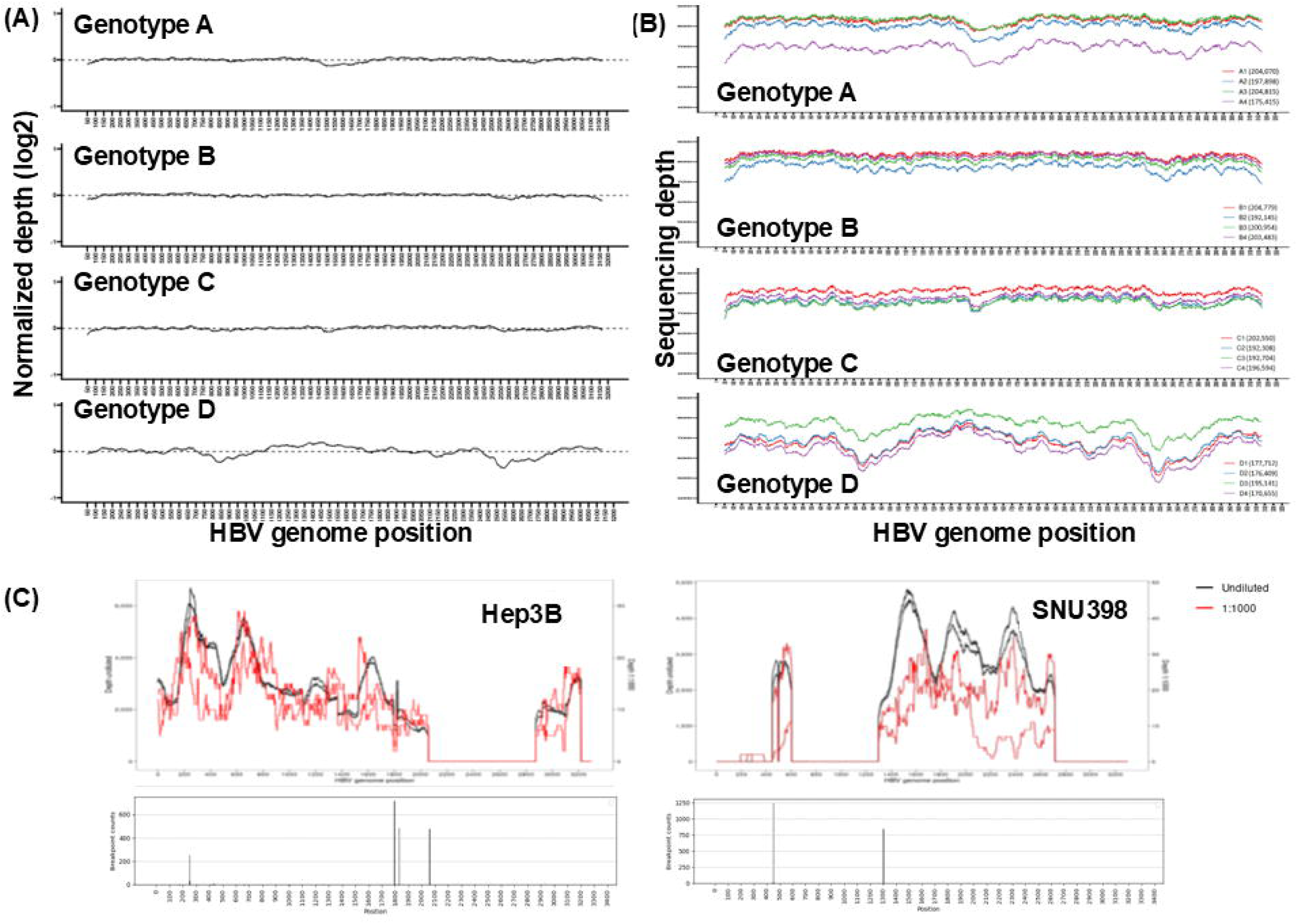
Characterization of the HBV-NGS assay. (A) HBV capture uniformity of the assay derived from the monomer of each of four genotypes, A, B, C, and D, with the normalized coverage depth from the mean (Y-axis) as shown for each nt position (X-axis) for each genotype. (B) Reproducibility of HBV-capture. Four independent experiments were performed using monomer of each of four genome types. The total UMI-consolidated HBV reads for each run are indicated in the parenthesis. X-axis indicates the position of the HBV genome position while the Y-axis indicates the coverage depths of each position. (C) The robustness (reproducibility and sensitivity) of HBV DNA capture. Two hepatoma cell lines, SNU398 and Hep3B, at undiluted (black lines) and 1:1000 dilution (red lines) in duplicates was subjected to HBV-NGS assay, as described in the text. HBV DNA coverage (top panel) and HBV-JS breakpoint (lower panel) maps are shown.

To assess the reproducibility of the assay, we performed the assay in replicates of four starting from the library construction using 1 ng of each linear monomer HBV DNA, followed by HBV panel hybridization and sequencing. As shown in **Figure 1B**, the captures of fragmented HBV DNA monomer templates from four genome types by the HBV probe panel demonstrates excellent reproducibility of each coverage pattern even though there were different by the total NGS reads, as indicated.

Next, we used DNA isolated from two HBV-HCC cell-lines that HBV DNA is only in the form of iDNA, Hep3B and SNU398, to assess the reproducibility. Two concentrations, 5 ng of undiluted HCC DNA and 15 ng of 1:1,000 dilution (0.2 ng of HCC DNA in 199.8 ng human DNA), as described in Material and Methods, were subjected to HBV-NGS assay and analyzed using the Advanced *ChimericSeq pipeline* for HBV-host junction identification and HBV DNA characterization. The HBV DNA coverage and HBV-host junction breakpoint maps were plotted (**Figure 1C**). As expected, the coverage map from both undiluted hepatoma cell line DNA overlapped in principle and to a lesser extent in 0.1% of both SNU398 and Hep3B, indicating that the HBV-NGS assay captured all HBV-JSs at 0.1% sensitivity. All detected junction viral breakpoint positions overlap in the plot, indicating excellent reproducibility even at 0.1%. This concludes that the HBV-NGS assay is a robust technology with an excellent capture uniformity of the HBV probe panel for the entire HBV genome, thus providing unbiased detection of HBV junction breakpoints with sensitivity of 0.1% at specificity at least 99.9% as none of the HBV DNA was detected when 30 ng of human DNA was tested as negative controls (data not shown).

### Sensitivity of iDNA detection

To demonstrate the HBV-NGS assay sensitivity of 0.1% is also applicable to detection of HBV-JS sequences. In addition to SNU398 and Hep3B which has only 2 [22] and 6 HBV-JS, respectively [23, 24], we included the PLC/PRF/5 cell line which has at least 10 HBV integrants and 15 known junction sequences, reported previously [19, 23–26]. Note that HBV DNA in these three hepatoma cell lines were reported as integrated partial HBV genomes. As a control, HepG2.2.15 cell line having stable viral expression and replication was included as a control for a full HBV DNA coverage for the assay.

To determine if HBV-NGS assay can detect all previously identified HBV-JS breakpoints at a sensitivity of 0.1%, we subjected 0.1% of diluted hepatoma cell line DNA with human genomic DNA and the undiluted HBV-HCC cell line DNA as positive controls.

As expected, HepG2.215 has full-HBV genome coverage by HBV-NGS assay (**Figure 2A**). In contrast to the other three HBV-HCC cell lines, only partial HBV DNA was detected as shown by HBV DNA coverage map. PLC/PRF/5 is known to contain at least 15 HBV junctions, as listed in **Figure 2B**, suggesting that at least 8 iDNA species exist in the cells. Interestingly, although SNU398 has been reported [22] to contain partial HBV DNA sequences, we discovered it contains only two detectable viral-host junctions at detection sensitivity of 0.1%, suggesting at least 99.9% of iDNA is from the same iDNA species, starting from HBV nucleotide position nt. 1,301 – 604 with recombination resulting in a 1,484 bp fragment deletion from nt. 2,719 – 447, and nt. 448 – 604 is inverted (**Supplementary 2A**). In addition to a 1,484 bp deletion, a 696 bp deletion (nt. 605 – 1,300) and a 3 nt deletion (nt. 497 – 499) were discovered, as the structure and the assembled full HBV iDNA sequence in SNU398 are summarized in **Supplementary Figure 2B and 2C**. Hep3B was reported to contain 4 – 6 junction sequences [23, 24]. The HBV-NGS assay is a short read NGS, thus limited in illustrating the structures of iDNA. We thus subjected Hep3B DNA to PacBio long read sequencing using methods previously described [24] and discovered two major iDNA species with one species at least two times more abundant than the other, consistent with HBV-JS NGS read counts from the HBV-NGS short read assay (**Supplementary Figure 3A**). Regardless, there is ∼800 bp common deletions for all iDNA resulting in the undetectable NGS reads from nt. 2,059 – 2,875 in HBV DNA coverage map at the sensitivity of at least 0.1% (**Supplementary Figure 3A**). The structures and the long read consensus sequences of these two major iDNA were shown in **Supplementary Figure 3B and C**. Consistently, all previously reported breakpoints, 15 from PLC/PRF/5, 6 from Hep3B, and 2 from SNU398, were detected by HBV-NGS assay at 0.1% sensitivity (**Figure 2B and C**).

**Figure 2.**
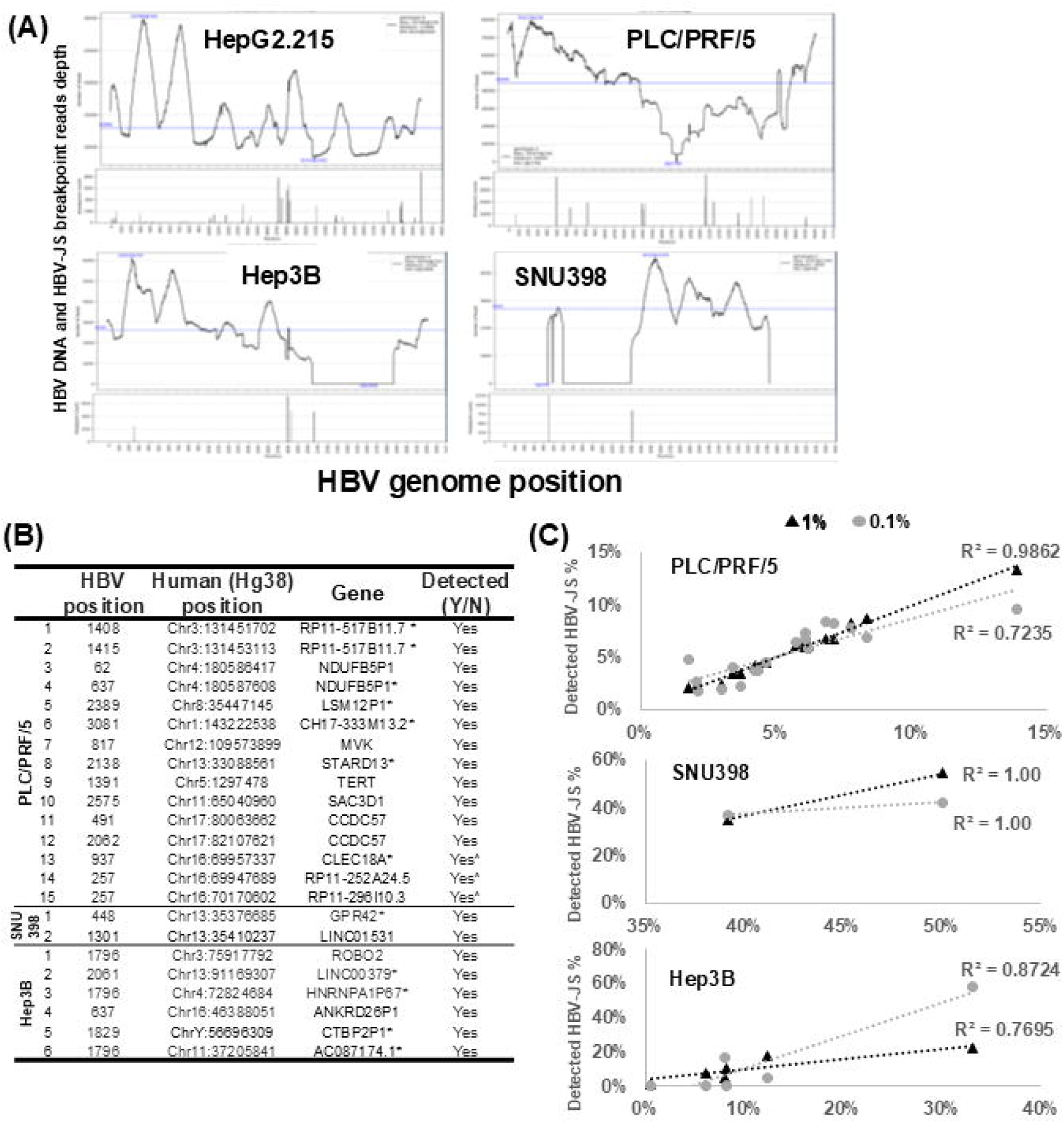
Sensitive detection and characterization of HBV-JS of HBV-HCC cell line DNA by HBV-NGS assay. **(A)** The HBV DNA coverage and HBV-JS breakpoint maps. The HBV genotype, maximum, minimum, and medium HBV read count are noted for each cell-line. **(B)** Detectability of previously reported iDNA species by HBV-NGS assay for PLC/PRF/5, SNU398, and Hep3B. *Breakpoint occurred in intergenic region and the closet annotated gene within 150 kb is listed; breakpoint detected at the other HBV junction end. **(C)** Assessment of HBV-JS sensitivity in reconstituted HBV-HCC cell-line DNA. Duplicates of each cell line DNA was reconstituted at 100%, 1%, and 0.1% in human DNA background and subjected to the HBV-NGS assay and sequenced at 20 million reads per sample except 100% samples which were sequenced at 10 million reads. For each individual HBV-JS, the average % HBV-JS of total HBV-JS reads was plotted for 1% (denoted in black triangle dot) and 0.1% (denoted in gray circle) samples against % HBV-JS detected in 100% samples. R^2^ is shown for each sample.

### Development of an iDNA estimation model using HBV-NGS assay

The HBV-NGS assay showed excellent uniformity and robustness for capturing HBV DNA across the entire genome. We hypothesized that the percentages of HBV-JS against total HBV DNA determined between using HBV-NGS assay and qPCR assays is positively correlated from patients when the majority of HBV DNA is iDNA, such as HBeAg (-) CHB patients with lower viral replication. If this is the case, we also hypothesized that an iDNA estimation model can be developed using quantities of HBV-JS of HBV DNA NGS reads in combination with iDNA sizes, if available.

To test these two hypotheses, biopsied tissue DNA obtained from four CHB patients with low viral loads (<4 log IU/mL) were subjected to HBV-targeted NGS assay to obtain HBV-JSs and HBV reads. HBV coverage and iDNA breakpoint maps are shown for each of the four patients in **Supplementary Figure 4**. The HBV-JSs were identified, and the percent of HBV-JS of total HBV reads were calculated for all junctions that had at least three supporting NGS reads. We next identified the most abundant HBV-JS from patients to test the hypothesis if a positive correlation can be established between HBV-NGS and qPCR assays for abundant HBV-JS. Junction specific PCR assays were designed and developed for quantification. DNA sequences of the junctions investigated, and the respective primers and PCR conditions are listed in **Supplementary Table 1**. A significant positive correlation was observed between % HBV-JS of HBV DNA determined by qPCR and by NGS using Pearson’s correlation test (r = 0.975, p < 0.001). We then determined if a linear model can be developed to estimate the percent of iDNA of the total HBV DNA by HBV-NGS assay in patient with low to undetectable viral replication, whose major HBV DNA species should be either covalently closed circular DNA (cccDNA) or iDNA.

A linear model for iDNA quantity prediction for each DNA sample was built by using % HBV-JS reads of total HBV reads by NGS and HBV-JS quantity per HBV DNA determined by CG1 qPCR from 6 abundant HBV-JS as detailed in Materials and Methods. In this model, as shown in **Figure 3**, the % HBV-JS calculated by NGS reads can explain 98% of the variance in the % HBV-JS calculated by qPCR (adjusted-R^2^ =0.98; p-value < 0.001) with a relationship of the qPCR values being 2.144× of the NGS values. The RMSE and R-squared value of this model obtained from LOOCV method were 8.74 and 0.91, respectively.

**Figure 3.**
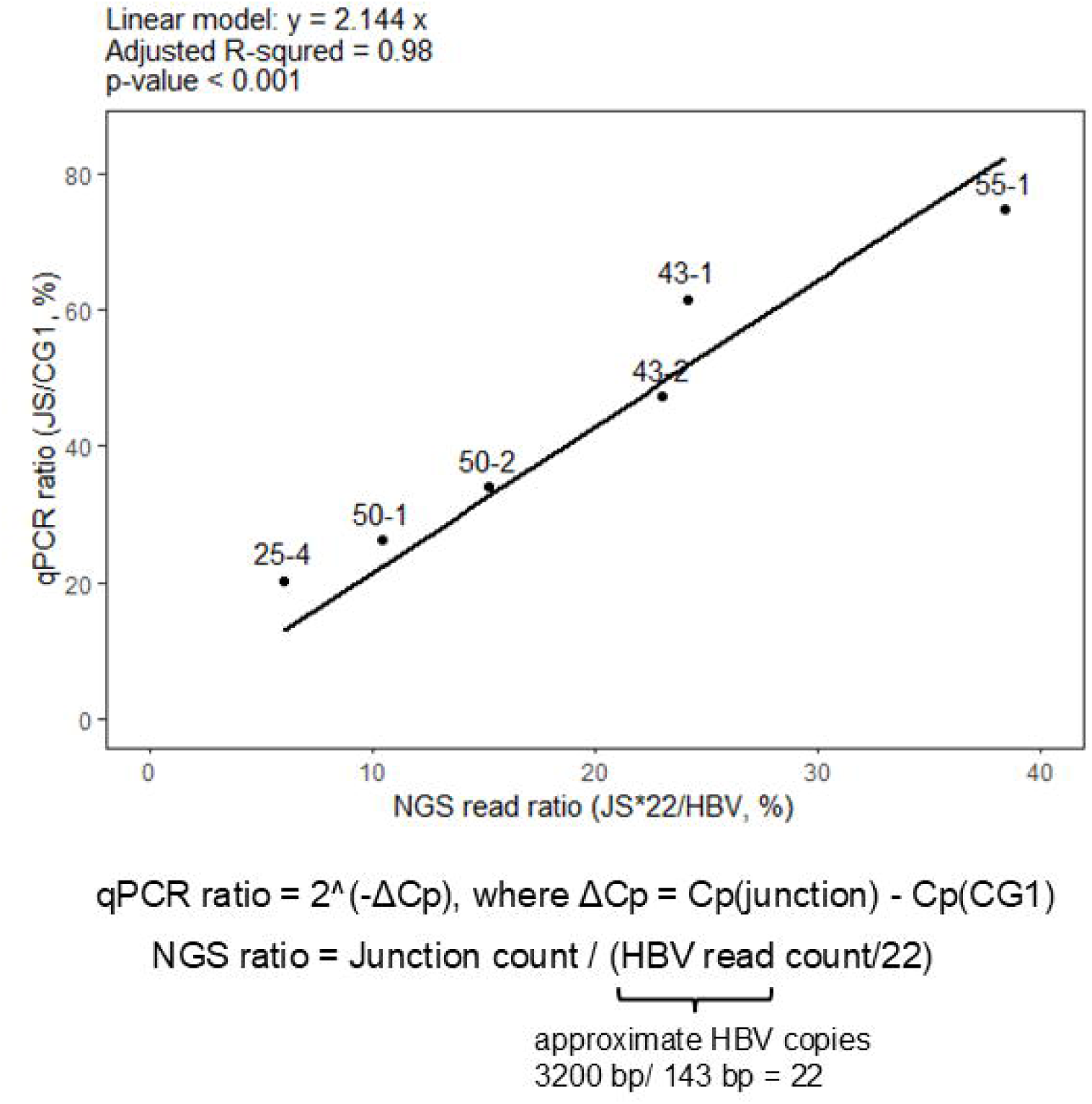
Development of a simple linear model for iDNA quantity estimation. Simple linear regression was used to find the association between junction proportions calculated from qPCR results and from NGS reads. The linear model was generated using function “lm” in R. The intercept was set to 0.

To validate the performance of this linear model for estimating the percent of iDNA of HBV DNA, we first used SNU398 DNA where all HBV DNA is iDNA and only one iDNA species exists as indicated by the detection of only two HBV-JSs, S1-GPR42 (hg38 Chr19:35376685, HBV nt. 448) and S2-LINC01531 (hg38 Chr19:35410237, HBV nt. 1,301) (**Figure 2**). The junction specific primers for the two junctions were designed and developed for quantitative measurement for each junction. As identified in **Figure 2A** and **Supplementary Figure 2**, SNU398 does not contain sequences coinciding with CG1 (nt. 246 – 309) so the HBV CG3 quantification assay (nt. 2,294 – 2,347) (Cat# qPCR-016, JBS Science) was used instead. By quantitating two junctions in SNU398 DNA by junction-specific qPCR assays, the ratio of S1/S2 is 1:1, as expected (**Supplementary Table 2**). Next, we determine the % HBV-JS by NGS using our developed model. We used iDNA size of 1.6 kb (as shown in **Supplementary Figure 2)** in our model for an equation as junction count*12/HBV read count, where 12 is the approximate counts of 143-bp HBV reads in a 1.6 kb iDNA. The ratio of the two HBV-JS is 1:1, even though the % HBV-JS between two HBV-JS are not 1:1 ratio in NGS data, as shown in **Figure 2A** by HBV-JS breakpoint plot, we estimated % iDNA based on the higher value of the % HBV-JS to accommodate the lower capture efficiency of the second HBV-JS. The iDNA was estimated as 89.0% and 97.0% from two independent HBV-NGS assays, an average of 93% of HBV DNA. Similarly, for determining the % HBV-JS in Hep3B, the higher % HBV-JS from each JS pair of the two major iDNA as described in the above section, was used to estimate %. Although we discovered all iDNA has at least 800 nt deletion by HBV DNA coverage map (**Figure 2**), two abundant iDNA sizes are 4.8 kb and 2.4 kb respectively. When we used ∼4.8 kb for iDNA #2 and 2.4 kb for the rest of iDNA for estimation, the iDNA estimation is approximately 83.0% and 84.4% from two independent HBV-NGS assays. As expected, two HBV-JSs of iDNA #1 are at ratio of 1:1 by the HBV-JS specific qPCR assays (**Supplementary Table 2**)

### Detection of HBV DNA and iDNA in liquid biopsy, plasma and urine, by HBV-NGS assay

The HBV-NGS assay has a high sensitivity of at least 0.1% suggesting its potential applications for HBV genetic liquid biopsy. Matched pairs of plasma and urine from three CHB patients with various serum viral loads, 6.3 log IU/mL (Pt A), <1 log IU/mL (Pt B) and undetectable (Pt C), were tested for potential applications of HBV genetic liquid biopsy using HBV-NGS assay. CfDNA (3 – 75 ng by availability) was subjected to NGS library construction and enriched for HBV DNA by two subsequent hybridizations except for Pt A which was by one run of hybridization, pooled and sequenced. NGS data was analyzed by Advanced *ChimericSeq pipeline*, plotted for HBV DNA coverage and junction breakpoint maps **(Figure 4A)** and summarized in **Figure 4B**.

**Figure 4.**
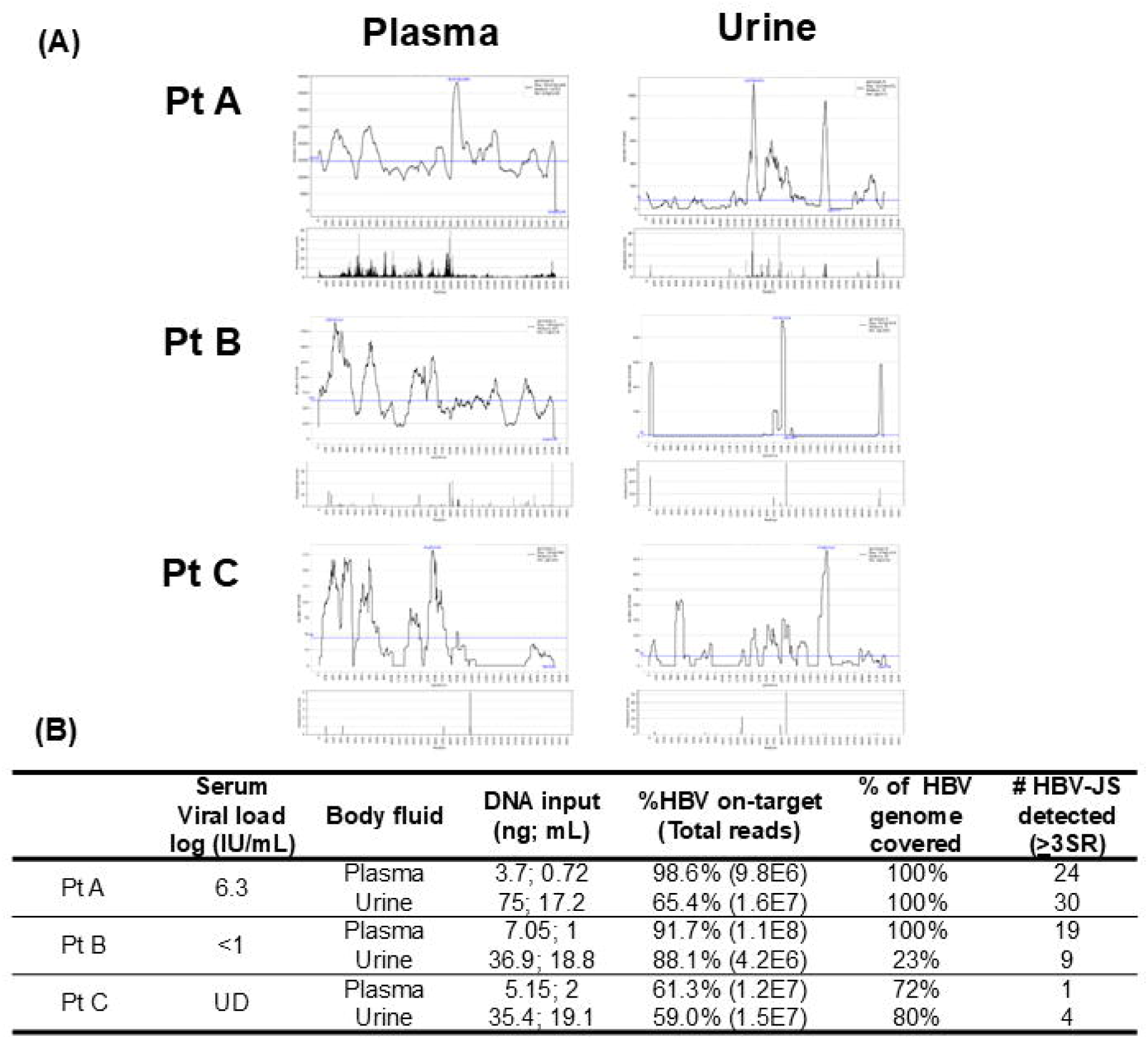
HBV genetic and iDNA liquid biopsy from patient with chronic hepatitis B infection. (A) The HBV DNA and junction breakpoint coverage of cell-free DNA isolated from matched plasma and urine, collected from three patients with serum viral loads from 6.3 IU/mL (Pt A), <1 IU/mL (Pt B), and undetectable (UD) IU/mL (Pt C). (B) HBV-NGS results.

As expected, the entire HBV genome sequence was detected from plasma collected from a patient with high viral load, Pt A. Overall, more HBV DNA can be detected from plasma than urine. Interestingly, from patient C with undetectable serum viral load, HBV DNA sequences can be captured and sequenced by HBV-NGS assay. A full genome sequence was obtained from Pt C, but only the partial genome was recovered from urine except for urine from high serum viral load CHB, Pt A. Encouragingly, HBV-JSs were also detected in all body fluid tested regardless serum viral loads.

## Discussion

This study applied a sensitive and robust hybridization capture assay of the entire HBV genome for NGS and used HBV-JS as a biomarker for iDNA to develop an iDNA estimation model using quantities of HBV-JS NGS reads in combination with iDNA sizes, if available for tissue biopsy from patients with limited or undetectable viral replication. The model was validated with two hepatoma cell-lines SNU398 and Hep3B. We demonstrated a sensitivity of 0.1% and specificity of at least 99.9% of iDNA detection (**Figure 1**) and detection of iDNA by liquid biopsy (from blood and urine) from the CHB patients with undetectable serum viral load. Furthermore, we reported, for the first time, the DNA sequences and rearrangement of iDNA of SNU398 cell-line and two major iDNA molecules of Hep3B cell-line.

We characterized an HBV-NGS assay and demonstrated that it uniformly enriches the HBV genome, enabling the possibility to estimate the iDNA fraction in total HBV DNA when HBV replication is limited. Although many iDNA studies have employed various techniques, those using high-throughput sequencing often do not to report key assay characteristics, particularly the uniformity or bias of HBV coverage and the sensitivity for detecting iDNA [3]. The robustness of an assay for iDNA detection is critical for interpreting findings. For example, the well-characterized HBV-HCC cell line PLC/PRF/5 is known to harbor at least seven HBV integration sites, yet different studies have reported varying numbers of integration sites [19, 23–26]. This discrepancy is likely due to differences in the assays used. Our HBV-NGS assay demonstrated high uniformity (within a 2-fold difference relative to mean coverage), reproducibility, and sensitivity of detection for iDNA, at least 0.1% (**Figure 1**). This precision allowed for the reliable estimation of the iDNA fraction in biopsied tissue DNA, using HBV-JS as a biomarker.

To develop a model for iDNA estimation, we considered several key factors. First, the model is intended for use in patients under effective antiviral therapy or with low viral loads, where most HBV species are either cccDNA or iDNA. Second, each iDNA molecule has two junction ends. Third, junction sequences have a disadvantage of being captured compared to full-length HBV DNA fragments. This is because junction sequences contain host DNA, which reduces their homologous complementary binding to HBV probes. Additionally, iDNA is often shorter than a full-length HBV DNA due to deletions introduced during the integration process. This could lead to an overestimation of iDNA if the model uses full-length sequences when iDNA sizes are not available as this is the case for patient tissue biopsy samples. However, it is possible that these two opposing factors may partially cancel each other out. To account for the unknown quantity of cccDNA that also contributes to the total HBV reads, we used a 3.2 kb (full-length HBV) reference to determine the relationship between junction fraction obtained from qPCR and those generated by HBV-NGS and to build the model to estimate the relative fraction of iDNA. The patients selected for model building had negligible viral loads, with full length HBV genome detected.

For model validation, we used the SNU398 and Hep3B cell lines, both of which are known to contain only iDNA. In the case of SNU398, HBV-NGS identified two junction sequences, suggesting the presence of a single iDNA species. By junction-specific qPCR assays, these two junctions were detected in a 1:1 ratio, as expected (**Supplementary Figure 2; Supplementary Table 2**). However, HBV-NGS analysis revealed that the S1 junction had significantly more reads than the S2 junction. To correct for the inefficiency in capturing junction sequences due to the presence of host DNA, we used S1 NGS reads (with a 1.6 kb iDNA size) to validate the model. This showed that, on average, 93% of the HBV DNA was iDNA in two independent HBV-NGS assays, thus validating the model for iDNA estimation. For Hep3B, which contains four major junctions, some minor junctions, and at least ∼800 bp of common deletions across all iDNA detected. We referenced the size of iDNA molecules using PacBio long-read sequencing. This allowed us to obtain two major iDNA sequences and their structural information. The two most abundant junctions, derived from the most prevalent iDNA species, were selected for the validation study. As expected, the two junctions were detected in a 1:1 ratio by junction-specific qPCR assays, with an estimated iDNA fraction of 83 – 84% (**Supplementary Figure 3; Supplementary Table 2**). Together, the estimation model is validated.

We have previously detected HBV DNA integration junctions in urine using a primer-extension capture based NGS approach [27]. HBV-NGS assay is more sensitive, at least 0.1%, a sensitivity that is applicable to circulating tumor DNA liquid biopsy. It was of interest to explore the application of HBV-NGS assay for iDNA liquid biopsy. Encouragingly, iDNA was detected in both blood and urine even from CHB patients with undetectable serum viral load.

There are several limitations to using the iDNA fraction estimation model. The estimation of the iDNA fraction is only applicable to the samples with limited viral replication. It would be difficult to estimate iDNA fraction in the samples with high level viral replication because of the high abundance and complexity of HBV DNA derived from viral replication. While the model has been validated using two hepatoma cell-lines when iDNA sizes are available, iDNA sizes are often not readily obtainable, thus limiting and complicating the estimation of iDNA fractions. Long-read sequencing, as demonstrated in the Hep3B study could aid in accuracy of the estimation. Ongoing research is incorporating factors like HBV DNA and HBV-JS breakpoint coverage maps and long-read NGS to further develop the model.

The model’s application to low-abundance junctions may be less accurate due to limitations in the data range (x-axis: 5.99 – 38.41%). As a result, the estimated iDNA percentage could be affected if samples contain a significant number of minor junctions. Although we attempted to broaden the data range by adding low-abundance junctions, these junctions were often difficult to quantify by qPCR, leading to limited data for model construction.

While the application of HBV-NGS for HBV genetic liquid biopsy is promising, as shown in this pilot study, a larger cohort is needed to fully assess its clinical potential. Nonetheless, this study highlights the robust analytical capabilities and high sensitivity of the HBV-NGS assay, providing an essential tool for investigating iDNA in HBV pathogenesis and disease management.

## Data Availability

All data produced in the present study are available upon reasonable request to the authors

## Acknowledgements

We thank Eli Xie for his assistance in performing qPCR testing for this study.

