## Supplementary Material for "Integrated DNA estimation in tissue biopsy and detection in liquid biopsy by HBV-targeted NGS assay"

**Authors’ affiliations:**

^1^ The Baruch S. Blumberg Research Institute, Doylestown, PA, 18902, USA

^2^ JBS Science, Inc., Doylestown, PA,18902, USA

^3^ Gilead Sciences, Inc, Foster City, CA, 94404, USA

^4^ Department of Microbiology and Immunology; Cancer Virology Program, UPMC Hillman Cancer Center, University of Pittsburgh School of Medicine, Pittsburgh, PA, 15213, USA

^5^ Liver Center, Department of Medicine, Beth Israel Deaconess Medical Center, Harvard Medical School, Boston, MA, 02215, USA

^6^ Department of Internal Medicine, National Cheng Kung University Medical College, Center of infectious Disease and Signaling Research, National Cheng Kung University, Tainan, Taiwan, Republic of China

**Corresponding Author:**

*Ying-Hsiu Su, The Baruch S. Blumberg Research Institute, 3805 Old Easton Rd, Doylestown, PA, 18902; Tel: 215-489-4949; Fax: 215-489-4920;.

### Supplementary Figures

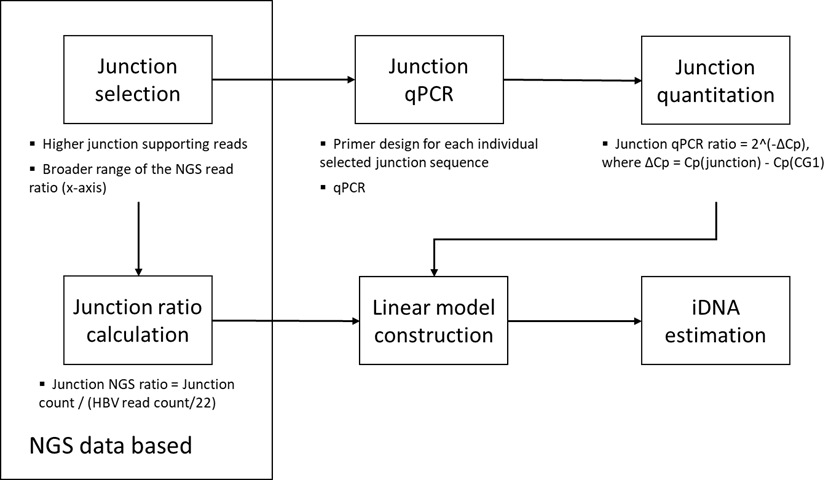

### Supplemental Figure 1. iDNA Estimation Model Development.

Development of a linear model for iDNA estimation by NGS reads for samples that have low viral load of <4 log IU/ml. This is under the assumption that there should only be limited viral replication in the low viral load group, thus the HBV DNA in infected liver should be either cccDNA or iDNA. In order to generate the linear model to estimate iDNA using NGS data, we identified 6 HBV-JS from low HBV viral load group that have abundances of at least 10% of total HBV-JS in that sample or can expand the applicable range (x-axis) of this model, to assess the model performance.

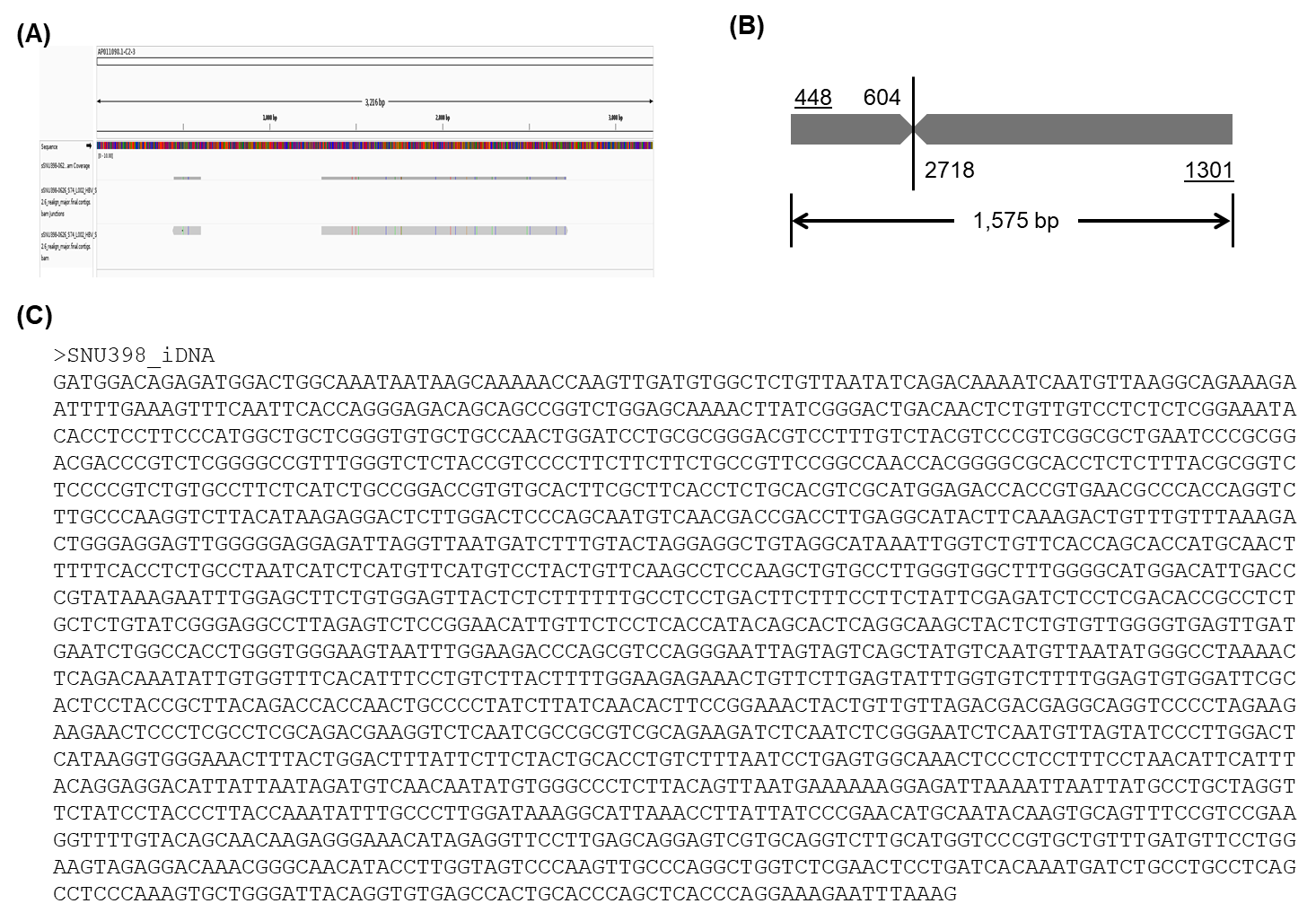

### Supplemental Figure 2. iDNA sequence in SNU398 cell-line DNA.

(A) IGV of HBV fragments from HBV-NGS. (B) iDNA structure. (C) iDNA sequence from sequence assembly.

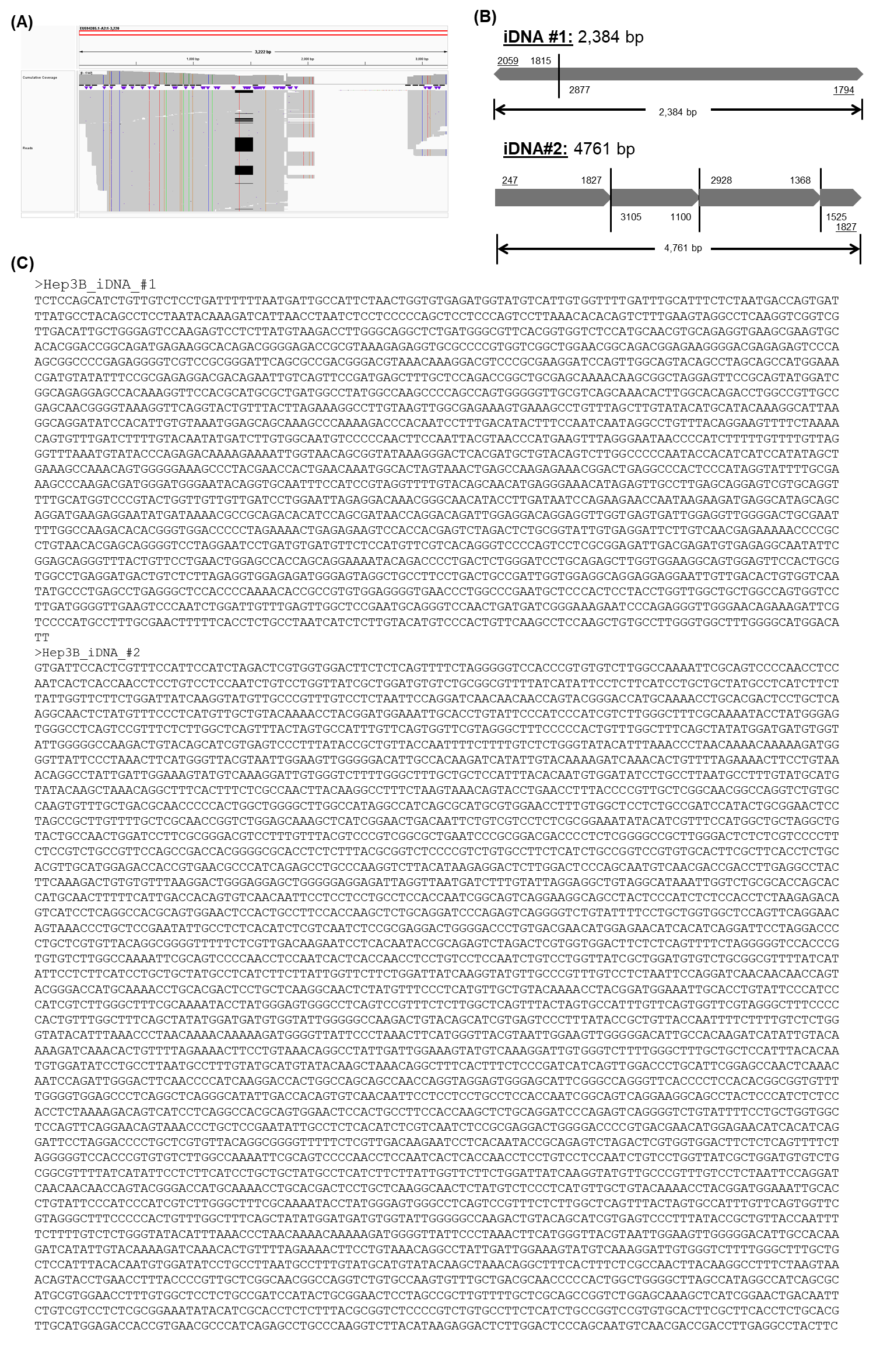

### Supplemental Figure 3. iDNA sequences in Hep3B cell-line DNA.

(A) IGV of HBV fragments from PacBio long reads. (B) iDNA structures. (C) iDNA sequences from long read consensus.

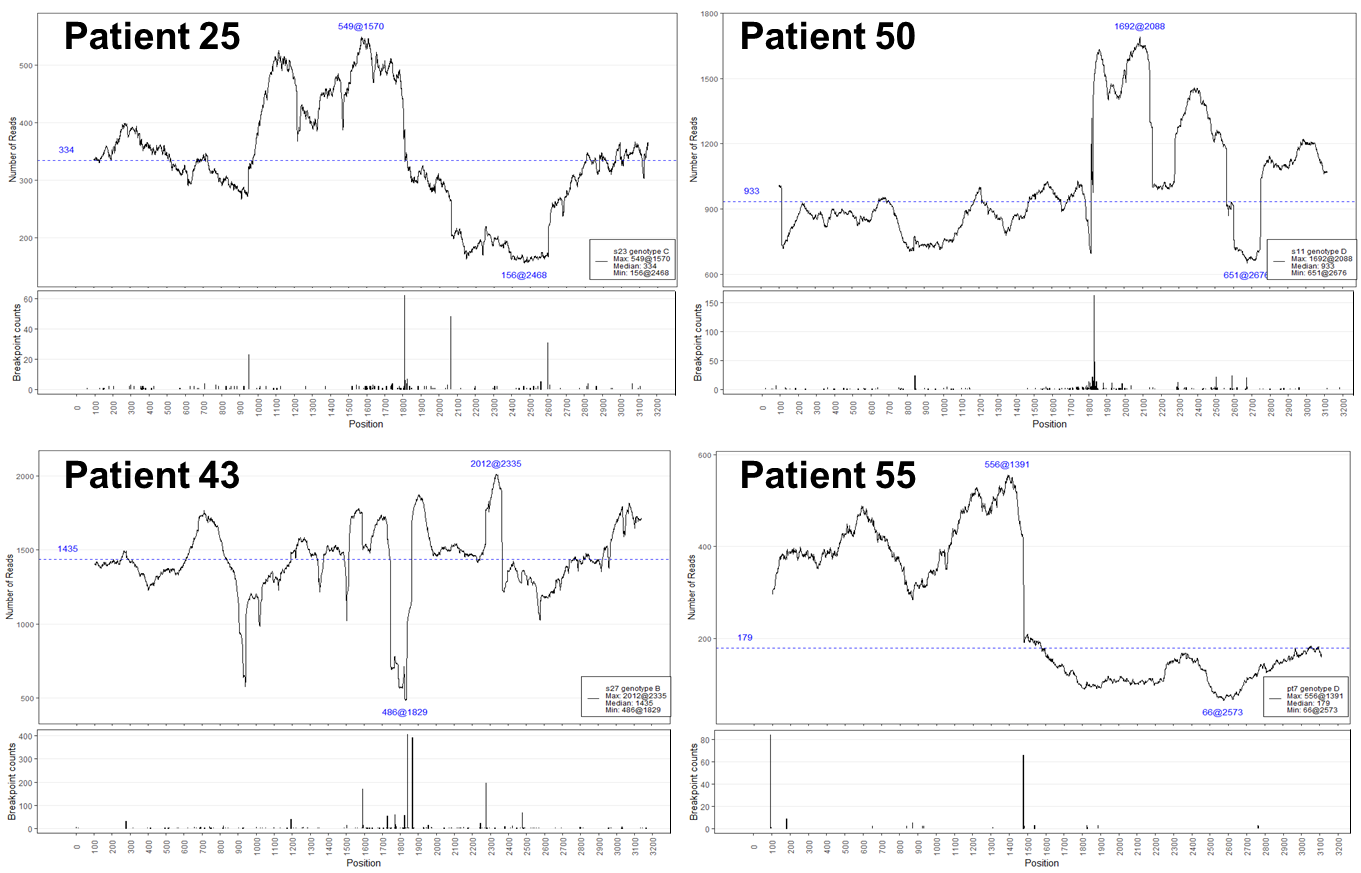

### Supplemental Figure 4. HBV DNA and iDNA breakpoint coverage maps of tissue DNA from four CHB patients.

### Supplementary Table

### Supplemental Table 1. Junction qPCR primers for iDNA estimation model development.

| **Patient ID** | **% HBV coverage** | **Junction ID** | **Junction coordinates**  **[hg38, HBV]** | **Junction SR (% in total HBV-JS)** | **Junction Sequence (5'-3')** | **Forward primer (5'-3')** | **Reverse primer (5'-3')** | **qPCR AT conditions** |
| --- | --- | --- | --- | --- | --- | --- | --- | --- |
| 55 | 100% | 55-1 | [Chr13:107773494, 94] | 84 (32.9%) | CAAAGCCTAAAAACGCATACTGTTGTTAAAAATAAGAGATCTTTGTAGAATAATACTTGGTTTTAGAAAAATTATTCAAAATACAGGAATAGAATTAGGACAAGAATAAGAAATCAGTCCGACTACTGCCTCTCACATATCGT | GATCTTTGTAGAATAATACTTGGTT | TATGTGAGAGGCAGTAGTCG | 60°C |
| 50 | 100% | 50-1 | [Chr8:5786566, 1829] | 125 (13.5%) | TAACTCCACAGTAGCTCCAAATTCTTTATAAGGGTCAATGTCCATGCCCCAAAGCCACCCAAGGCACAGCTTGGAGGCTTGAACAGTGGGACATGTACAAGAGATGATTAGGCAGAGCTGGGTGGCGTGGAGCATGGGATGAAGCAAGCAACACGAAGCTGTCATTCCCAGCCTCTGTGTGTCATTCTTAGATAATTTTGCCCCAGTACACTGCTGCATCTTCTCAACTGGACTT | CAGTGGGACATGTACAAGAGAT | TTGAGAAGATGCAGCAGTGTA |  |
|  |  | 50-2 | [Chr8:5786557, 1831] | 182 (19.6%) | AGAGTAACTCCACAGTAGCTCCAAATTCTTTATAAGGATCAATGTCCATGCCCCAAAGCCACCCAAGGCACAGCTTGGAGGCTTGAACAGTAGGACATGAACAAGAGATGATTAGGCAGGACTGCACTGGTCCTGCGAGCTCCCCTCCTGGCGCACAGGAGAGCAGAGGGGCCACCATTTGCCTGGCTGTCGGGCTGTCCCTGAAGAACTGCTTTAGTTTCTTCCTGTCGAGATTGC | GCTTGGAGGCTTGAACAGTA | CTAAAGCAGTTCTTCAGGGACA |  |
| 25 | 100% | 25-4 | [Chr4:77818365, 950] | 23 (5.79%) | GCATATAAAGGCATTAAGGCAGGATAGCCACATTGTGTAAAAGGGGCAGCAAAGCCCAAAAGACCCACAATTCTCTGACATACTTTCCAATCAATAGGTCTATTTACAGGCAGTTTTCACAAAATCCATCCAATCCCACAGGCCAGGAAAGCCAGTTTCCCTTTCTTCCTCAGCCCTTCCAAGAAAATGTTTGTCTAATTAAAAAAAAAAAAATCTTACNNACG | TAAGGCAGGATAGCCACATTG | GCTGAGGAAGAAAGGGAAACT |  |
| 43 | 100% | 43-1 | [Chr5:57298932, 1839] | 408 (21.0%) | CAAAAAAGAGAGTAACTCCACAGAAGCTCCAAATTCTTTATACGGGTCAATGTCCATGCCCCAAAGCCACCCAAGGCACAGCTTGGAGGCTTGAACAGTAGGACATGAACATGAGATGACAGAGCCCCTGTATGACAGTTGTTTAGAACAGAAACATGCTCTTTTTGCCTCTCCTCCAGCCACCACCCACTCCACTACCATGATCAGTAAGGTGGGAAGGAATCCAAAGGGCCATAG | TATACGGGTCAATGTCCATGC | CTTACTGATCATGGTAGTGGAGTG |  |
|  |  | 43-2 | [Chr5:57298966, 1867] | 389 (20.1%) | TATTAAAAAAGTTTAACACACCCTGATTAGTCTCTTTCCTTTTCTTCTCAGCCTCTCCAAAGAGAATCATCTCTTGACTGTCACTGCAATAGCCAAATTAAACCACAAATTAACCCACAGCCTCCAAGCTGTGCCTTGGGTGGCTTTGGGGCATGGACATTGACCCGTATAAAGAATTTGGAGCTTCTGTGGAGTTACTCTCTTTTTTGCCTTCTGACTTCTTTCCTTCT | CTCAGCCTCTCCAAAGAGAATC | AACTCCACAGAAGCTCCAAAT |  |
| SNU398 | 49% | S1 | [Chr19:35376685,448] | 1234 (59.1%); 1436 (60.7%) | ATAGAGGTTCCTTGAGCAGGAGTCGTGCAGGTCTTGCATGGTCCCGTGCTGTTTGATGTTCCTGGAAGTAGAGGACAAACGGGCAACATACCTTGGTAGTCCCAAGTTGCCCAGGCTGGTCTCGAACTCCTGATCACAAA | GCAACATACCTTGGTAGTCC | GATCAGGAGTTCGAGACCAG | 60°C |
|  |  | S2 | [Chr19: 35410237,1301] | 852 (40.8%); 923 (39%) | AAAAACCAAGTTGATGTGGCTCTGTTAAAATCAGACAAAATCAATGTTAAGGCAGAAAGAATTTTGAAAGTTTCAATTCACCAGGGAGACAGCAGCCGGTCTGGAGCAAAACTTATCGGGACTGACAACTCTGT | AAGTTTCAATTCACCAGGGAGACAG | AGTCCCGATAAGTTTTGCTCCA | 62°C |
| Hep3B | 75% | H1 | [Chr13:91169310,2056] | 784 (33.6%); 810 (36.2%) | ACACCGCCTCAGCTCTGTATCGGGAAGCCTTAGAGTCTCCTGAGCATTGCTCTCCTCACCATCATTGGTAGACAGAGAGTCTGATAGGAGTTGTTAGGAAACAGACAGAGAGAGCAAATATCTGGAAACCCT | CCTTAGAGTCTCCTGAGCATTG | TCCTATCAGACTCTCTGTCTACC | 55°C |
|  |  | H2 | [Chr3: 75917792,1793] | 698 (30%); 687 (30.7%) | GTGTGTTTAAGGACTGGGAGGAGCTGGGGCGGAGATTAGGTTACTGATCTTTGTATTAGGAGGCTTTAGGCCTAAATCACTGGTCATTAGAGAAATGCAAATCAAACCCACAATGACATACCATCTCACACCA | GATTTGCATTTCTCTAATGACCAGTG | CTTTGTATTAGGAGGCTGTAGGC | 62°C |

AT, denotes specific qPCR assay annealing Tm where each qPCR protocol is as follows “95°C, 5 min, 45 cycles (95°C, 10 s, AT °C, 10 s 72°C, 10 s)”

Note: underlines indicate HBV sequences.

### Supplemental Table 2. iDNA junction ratio in SNU398 and Hep3B cell-line DNA.

| Average Cq ± SD | **S1** | **S2** | **CG3** |
| --- | --- | --- | --- |
| SNU398 | 25.50 ± 0 | 25.19 ± 0.03 | 25.13 ± 0.01 |
|  | 0.81 (S1/S2 ratio)* | |  |
| Average Cq ± SD | **H1** | **H2^#^** | **CG1** |
| Hep3B | 26.23 ± 0.153 | 26.04 ± 0.017 | 27.67 ± 0.007 |
|  | 0.876 (H1/H2 ratio* | |  |

*Calculated as described in the Methods using the formula: 2^(-ΔCp); ^#^, Cq values were normalized by amplification efficiency between H1 and H2 qPCR assays.
